# Shifts in Play Experiences and Impact on Preschool Children’s Development During COVID-19

**DOI:** 10.1101/2025.10.10.25337686

**Authors:** Nigar Sultana, Maliha Mahazabin

**Affiliations:** Department of Child Development and Social Relationship, College of Home Economics, Azimpur, Dhaka 1205, Bangladesh; Special Education Teacher, Canadian Trillinium School, Gulshan – 2, Dhaka; Statistics Discipline, Science Engineering and Technology School, Khulna University, Khulna – 9208, Bangladesh

**Keywords:** Preschool children, COVID-19 pandemic, Family support, Play, Child development, Bangladesh

## Abstract

The COVID-19 pandemic and prolonged lockdowns severely disrupted opportunities for play among preschool children. Play is a vital aspect of early childhood, fostering both physical and mental development, as active play strengthens physical ability and supports emotional, social, and cognitive growth. This qualitative study examined the consequences of restricted play during the pandemic on children’s health and development in Bangladesh. Results indicated that limited opportunities for active play adversely affected both physical and mental well-being, with mental health being more significantly compromised. Commonly reported issues included overall decline in health, irritability, increased reliance on electronic devices, irregular sleep patterns, unhealthy dietary changes, delays in language development, and reduced socialization. Parents and guardians faced substantial challenges in addressing their children’s play needs. The greatest difficulty was restricting outdoor play, while simultaneously attempting to create safe indoor play arrangements, reorganize household space, and introduce new activities. Despite these efforts, many were unable to meet children’s developmental needs within the confines of the home. These findings underscore the essential role of play in early childhood and highlight the importance of developing supportive strategies to protect child development during future public health emergencies.

## 1. Introduction

Play is a spontaneous and instinctive activity for children, essential to their growth, learning, and overall well-being. It is not merely a way to pass time or keep children occupied, but a fundamental component of healthy development. Friedrich Froebel, the founder of the kindergarten movement, described play as “the purest, most spiritual activity of man at this stage … typical of human life as a whole” **(Fröbel, 1885)**. Herbert Spencer’s surplus-energy theory (1870s) suggested that play is a way for children to expend energy that is not required for survival or basic tasks **(Spencer, 1870)**, while Jean Piaget viewed play—especially pretend and symbolic play—as important for cognitive development, enabling children to experiment, assimilate new experiences, and interact socially, which supports the construction of intelligence **(Piaget, 1951)**. Lev Vygotsky (1978) further argued that play is a critical context in which children develop higher mental functions and social competence **(Vygotsky & Cole, 1978)**. Together, these theories underline the importance of play for the physical, cognitive, social, and emotional development of young children.

Play contributes to physical health by strengthening muscles, bones, and motor coordination, while also stimulating creativity, imagination, and problem-solving skills. It enhances social relationships, emotional regulation, and resilience **(Ginsburg et al., 2007; Yogman et al., 2018)**. Importantly, the nature of play evolves with age, becoming more complex and diverse as children grow, but its developmental value remains constant. The COVID-19 pandemic, however, drastically disrupted children’s play opportunities worldwide. Prolonged lockdowns, school closures, and restrictions on outdoor activities limited children’s ability to engage in free and social play **(UNICEF, 2022)**. In Bangladesh, as elsewhere, preschool children were confined to their homes for extended periods, with limited alternatives for play and interaction. This isolation restricted peer relationships, disrupted routines, and negatively affected both physical and mental health. Parents attempted to create indoor play arrangements and reorganize household spaces, but these efforts were often insufficient to meet children’s developmental needs. Many children experienced loneliness, irritability, irregular sleep, unhealthy dietary habits, dependence on electronic devices, delayed language development, and reduced opportunities for socialization **(Lee et al., 2024; Singh et al., 2020)**.

The COVID-19 pandemic significantly disrupted children’s play and development worldwide. Alcon et al. conducted a systematic review and reported that preschool children experienced negative impacts on both mental and physical development due to reduced play opportunities **(Alcon et al., 2024)**. Paterson et al. highlighted increased sedentary behavior, reduced physical activity, and altered sleep patterns among children and youth during lockdowns **(Paterson et al., 2021)**. Prior study found that children exhibited higher rates of anxiety, depression, and behavioral problems, emphasizing the pandemic’s adverse effects on mental health **(Ng & Ng, 2022)**. McIsaac et al. observed that while families implemented indoor play adaptations, these strategies often could not fully compensate for lack of peer interaction and outdoor play opportunities **(McIsaac et al., 2024)**. In Bangladesh, similar patterns were documented, with school closures and home confinement leading to disruptions in routines, socialization, and early learning, and increasing parental stress **(Singh et al., 2020)**. Collectively, these studies indicate that restricted play during COVID-19 negatively affected children’s physical, mental, and social development, underscoring the importance of strategies to support play during crises.

Children were particularly vulnerable because they could not fully understand the reasons for restrictions or the implications of the virus, making the sudden changes in daily routines confusing and emotionally stressful. Globally, the pandemic has been recognized as disproportionately affecting vulnerable populations, including the elderly and young children **(Loades et al., 2020)**. For preschoolers in Bangladesh, these challenges translated into significant disruptions to play, learning, and healthy development. The aim of this study is to explore the impact of COVID-19 lockdowns on preschool children’s play in Bangladesh. Specifically, it investigates: (1) the types of play children engaged in during the pandemic; (2) the play activities they preferred and the time spent on them; (3) the extent of opportunities available for play; (4) the facilities and strategies adopted by families to support play; (5) the effects of restricted play on children’s physical and mental health; and (6) the challenges parents faced in arranging play during the lockdown.

## 2. Materials and methods

### Study Design and Approach

This study employed a qualitative research design to explore the impact of COVID-19 on preschool children’s play and development in Dhaka, Bangladesh. Qualitative methods were chosen to obtain in-depth insights into parents’ experiences, perceptions, and strategies regarding their children’s play during the pandemic. This design provided a deeper understanding of how families adapted to challenges and how restricted play affected children’s developmental outcomes.

### Study Area

The survey was conducted in selected urban areas of Dhaka city, including Azimpur, Uttara, Shahabag, and Dhanmondi, chosen to capture diverse socio-economic contexts and living environments that could influence children’s access to play opportunities during the lockdown.

### Participants and Sampling

Participants were mothers of preschool children aged 2–3 years. Purposive sampling was employed to select participants who were directly responsible for their children’s care and could provide detailed accounts of play practices and challenges during the pandemic. The distribution of participants is summarized in Table 1.

**Table 1.** Participant Distribution by Area, Mother Age and Number of Children.

| Area | Mother Age (years) | Number of Children |
| --- | --- | --- |
| Azimpur | 26-30 | 2 |
| Azimpur | 26-30 | 1 |
| Azimpur | 26-30 | 1 |
| Uttara | 26-30 | 1 |
| Uttara | 31-35 | 2 |
| Uttara | 26-30 | 1 |
| Shahabag | 26-30 | 2 |
| Shahabag | 26-30 | 2 |
| Dhanmondi | 26-30 | 1 |
| Dhanmondi | 31-35 | 1 |

### Data Collection

Semi-structured interviews were conducted with the participants. Prior to the interviews, the study objectives were clearly explained, and written informed consent was obtained from mothers. Interviews focused on children’s play activities, parental strategies to support play, time allocation, impacts on physical and mental health, and challenges faced during the COVID-19 lockdown. Each interview lasted approximately 30 minutes and was conducted face to face between 3^rd^ February to 25^th^ April 2022.

### Ethical Considerations

Participation was voluntary, and participants were informed of their right to withdraw at any time. Data were anonymized to ensure confidentiality, and ethical clearance was obtained from the relevant institutional review board prior to data collection.

### Data Analysis

Data were analyzed thematically following the six-phase framework proposed by Braun and Clarke **(Braun & Clarke, 2008)**, which involved familiarization, coding, theme development, review, definition, and interpretation. This approach allowed for identifying recurring patterns and meanings in participants’ experiences. Key themes regarding children’s play patterns, parental strategies were identified, coded, and cross-verified among the research team to enhance reliability.

## 3. Results

The study explored how COVID-19 lockdowns affected preschool children’s play activities and overall development in Bangladesh. Analysis of parent interviews revealed several key themes related to children’s daily play, family relationships, behavioral changes, and physical and psychological impacts.

### Outdoor Play

During lockdown, most children did not have the opportunity to play outdoors. Parents were cautious about allowing their children to meet others outside due to safety concerns. Some parents occasionally took their children outdoors when it seemed safe, while others allowed visits to nearby relatives’ houses so children could meet and play with cousins. A few children who had open spaces around their homes were taken to rooftops, front yards, or empty spaces nearby for short periods of play while maintaining hygiene measures.

However, some children were not allowed outdoors at all because of strict parental rules. Despite these restrictions, most parents expressed a strong desire to provide their children with at least limited outdoor play opportunities, even if only for short periods.

### Kinship or Relationship with Family Members

During COVID-19, families spent long hours indoors, which strengthened interpersonal relationships. Children living in extended families developed closer bonds as family members had more time to spend together. Many parents who were home from work reported that they could give more attention to their children, creating a joyful and engaging home environment. In nuclear families, the situation varied. Some parents were able to spend more quality time with their children than before the pandemic, reducing loneliness and improving family bonding. However, other parents, especially those working from home, found it difficult to balance office work and household duties. As a result, some children felt lonely, and their bonding with parents was negatively affected.

### Indoor Playing Environment

When outdoor play was restricted, children fulfilled their need for play through different indoor activities. Many children played with their siblings or family members, while others played alone using available toys or creative materials. Examples of indoor play included block building, drawing, painting, playing with paper, or water play. Some parents even bought small plastic slides for their children so they could enjoy playtime indoors. Overall, parents made considerable efforts to create enjoyable and engaging indoor play environments for their children.

### Children’s Play Preferences and Time Spent

Many parents mentioned that their children preferred more active and social play, but indoor space and limited equipment restricted those options. As a result, children often played in short intervals and shifted quickly between activities. Parents noted that children’s attention span for play decreased because of repeated use of the same toys and lack of variety. These finding highlights that both children’s play preferences and the duration of playtime were strongly influenced by environmental constraints during lockdown.

### Affection to Electronic Devices

Because children were confined indoors, many gradually lost interests in traditional toys after long-term use. At that stage, several parents allowed their children to use electronic devices such as smartphones or tablets to keep them entertained. Over time, some children became highly addicted to these devices, spending long hours playing digital games or watching videos. When this addiction grew, some parents tried to reduce screen time by introducing new toys or spending more time with their children. A few children, however, preferred constructive and creative activities rather than electronic devices during the lockdown.

### Language Conflict

During lockdown, children were often exposed to cartoons and animation videos in foreign languages, especially English, as a major source of entertainment. As a result, many children became more comfortable understanding and responding in the foreign language than in Bangla, which is used for family communication. This situation created a form of language conflict—children sometimes struggled to express their needs or emotions clearly in Bangla, leading to communication difficulties between them and their parents. Consequently, some children became less interested in verbal communication.

### Food Habits

Reduced outdoor play and physical activity, along with a monotonous lifestyle, negatively affected children’s eating habits. Parents noticed decreased appetite and aversion to food among their children. To keep them happy and distracted from outdoor cravings, parents often provided foods that children found attractive, such as chips, chocolates, and fast food. As a result, maintaining a balanced and nutritious diet during the lockdown became difficult.

### Sleep

Children who are physically active tend to have better sleep patterns. During COVID-19, limited opportunities for play reduced children’s physical activity, leading to irregular sleep habits. Many parents reported that their children stayed awake late at night and woke up late in the morning. Sleep deprivation made children more irritable, easily frustrated, and prone to tantrums.

### Physical Effect

The lockdown period had several negative effects on children’s physical development. Due to the lack of outdoor play such as running, cycling, and climbing, children’s large muscle movement and motor skill development were hindered. Some children who could previously ride bicycles or climb before the pandemic forgot these skills after long confinement at home. In addition, changes in diet and reduced physical activity affected their immunity, leading to frequent illness. Some children became overweight due to inactivity, while others showed reluctance to participate in active games. Increased screen uses also caused eye problems such as redness and watery eyes

### Psychological Impact

Lack of play opportunities acted as a barrier to children’s balanced mental development. The prolonged home confinement and limited social interaction created stress and frustration. Parents reported behavioral problems such as stubbornness, excessive anger, irritability, and disobedience. Some children displayed aggressive or destructive behaviors—shouting, hitting family members, pinching, throwing or breaking objects, and damaging toys. These behaviors reflected emotional distress and frustration resulting from a lack of active play and socialization.

### Creativity

Despite the challenges, many parents encouraged creative activities to keep their children engaged. They provided materials like painting books, picture books, and colored pencils. Parents also read storybooks or told stories aloud, which enhanced children’s imagination and creative thinking. Children engaged in constructive play by using everyday household materials to create new objects. For example, they pretended to cook or make items using vegetables, flour, rice, or lentils. These activities helped maintain creativity and curiosity during the lockdown.

### Challenges Faced by Parents

Parents faced multiple challenges while arranging play activities during the lockdown. The biggest difficulty was preventing children from going outdoors while still keeping them active and happy indoors. Limited indoor space, lack of appropriate toys, and balancing household and work responsibilities made it difficult for parents to consistently organize play sessions. Many also struggled to replace the social aspect of play that children experienced with peers. Despite these obstacles, parents tried their best to create a playful atmosphere within the home using available resources.

### Impact After Lockdown

When restrictions were lifted, changes were observed in how children adjusted to the return of normal play routines. Some children preferred solitary play and avoided social interaction with peers, suggesting that long periods of isolation affected their social development. Others expressed great excitement and joy in being able to go outside and play with friends again. These mixed responses indicate that the lockdown had lasting but varied effects on children’s social behavior and play patterns.

## 4. Discussion

The findings of this qualitative study reveal that preschool children experienced significant disruption to both physical and mental development due to prolonged home confinement during the COVID-19 pandemic. The guardians reported that children’s opportunities for active play were drastically reduced, leading to a noticeable decline in large muscle movement, physical stamina, and overall health. Several participants mentioned emerging health concerns, including obesity, weakened immunity, and general physical inactivity. These outcomes are consistent with findings from earlier study observed that children worldwide experienced declines in physical activity levels and increases in sedentary behavior and irregular sleep patterns during the pandemic **(Paterson et al., 2021)**. Such restrictions on outdoor movement and active play directly limited the development of motor skills, which are critical during early childhood.

The study also identified notable behavioral and emotional challenges among preschool children. Guardians frequently reported irritability, excessive anger, stubbornness, aggression, hyperreactivity, and reduced alertness in their children. Many children also appeared more withdrawn, sad, or less enthusiastic about daily activities. These findings align closely with a prior study reported that the pandemic period led to an increase in emotional distress, anxiety, and depressive symptoms among children due to social isolation and reduced peer interaction **(Ng & Ng, 2022)**. Similarly, Alcon et al. found that disruptions in play, routine, and social connection during COVID-19 were associated with delays in both cognitive and socio-emotional development in early childhood populations **(Alcon et al., 2024)**. Moreover, behavioral changes like irritability, aggression, and depression observed in our study are corroborated by **Loades et al., 2020**, who conducted a rapid systematic review and found that social isolation and loneliness during the pandemic significantly increased the risk of depression and anxiety in children and adolescents. The results of this study further support **McIsaac et al., 2024**, who described how the nature of children’s play shifted significantly during the pandemic. With limited access to playgrounds and outdoor spaces, parents attempted to create alternative play opportunities indoors; however, these often failed to meet children’s developmental needs. In the present study, guardians expressed similar difficulties, stating that even though they tried to engage children in home-based play, the lack of physical space and social interaction hindered meaningful play experiences. This underscores how environmental and contextual factors shape the quality of play, which in turn affects physical and psychological development.

The observed decline in physical activity among children during lockdowns is consistent with prior findings which revealed that pandemic-related restrictions led to reduced physical activity in children and adolescents, contributing to various health issues such as obesity and weakened immunity **(Singh et al., 2020)**. Our findings regarding parents’ struggles to provide adequate play opportunities aligns studies which highlighted that caregivers faced challenges in maintaining children’s mental health during lockdowns due to limited resources and increased stress **(Lee et al., 2024; Ng & Ng, 2022; Singh et al., 2020)**. Collectively, the results highlight the multidimensional consequences of prolonged home confinement on preschool children in Bangladesh, echoing international findings. The combination of reduced physical activity, social isolation, and screen dependency likely amplified both physical and mental health problems among young children. Moreover, the findings suggest that parents faced considerable challenges in balancing safety concerns with the need for play and socialization, a dilemma similarly documented in other global contexts **(Alcon et al., 2024; McIsaac et al., 2024)**. These findings emphasize the importance of developing child-friendly public health responses during emergencies. Policies should ensure that children have safe and accessible opportunities for physical activity and social play, even under movement restrictions. Additionally, parental support programs are essential to help families manage children’s emotional well-being and promote adaptive play strategies during crises.

This study has several limitations. First, it involved a small sample of guardians from selected urban areas of Dhaka, which limits the generalizability of the findings to all preschool children in Bangladesh. Second, as a qualitative study, the data relied entirely on parent reports, which may be influenced by recall bias or subjective interpretation of children’s behaviors. Third, the study focused on a specific pandemic context, so results may not fully apply to other crises or situations with different restrictions. Despite these limitations, the findings provide important insights into the impact of prolonged home confinement on preschool children’s physical and mental development. Future research should consider larger and more diverse samples across urban and rural areas, incorporate longitudinal designs to track long-term effects, and include direct observation or child-reported data to complement parental perspectives. Such studies could strengthen understanding of how play interventions and home environments can mitigate developmental challenges during pandemics or other emergency situations.

## 5. Conclusion

This study highlights that preschool children in Bangladesh experienced significant disruptions in both physical and mental development during the COVID-19 lockdown due to limited play opportunities. Most children developed strong attachment to electronic devices, while only a few engaged in creative or constructive play. Irregular sleep patterns, deteriorated food habits, and language development difficulties were widely reported. Some children also lost interest in outdoor play.

Overall, restricted play during the pandemic negatively affected children’s development, with mental health being more profoundly impacted than physical health. Guardians faced considerable challenges in arranging play, particularly in preventing children from going outdoors while attempting to meet their developmental and recreational needs. Despite these difficulties, parents made considerable efforts to provide stimulating and enjoyable play environments within the home.

### Recommendations

Based on the findings, the following measures are recommended to support preschool children’s development during future pandemics or similar emergencies:

- **Ensure safe opportunities for play during restrictions**: Policymakers and caregivers should facilitate safe indoor and limited outdoor play options to maintain physical activity and social interaction even during lockdowns.
- **Minimize excessive screen time**: Structured schedules and engaging alternatives should be provided to prevent overreliance on electronic devices and encourage creativity.
- **Promote physical and motor skill activities**: Parents and caregivers should be guided to incorporate simple, age-appropriate exercises at home to maintain children’s physical health, large muscle development, and sleep quality.
- **Parental guidance and support programs**: Resources and training for parents can help manage behavioral challenges, support mental health, and create stimulating indoor environments.
- **Integrate crisis-responsive early childhood education strategies**: Educational authorities should develop play-based interventions that are adaptable during emergencies to ensure cognitive, social, and emotional development continues despite restrictions.

These recommendations aim to prepare families, educators, and policymakers to safeguard preschool children’s holistic development during any future pandemic or emergency situations where normal play routines are disrupted.

## Author Declarations

This work did not receive any fund. The authors declared no competing interest.

All relevant ethical guidelines have been followed, the study received approval Ethical Clearance Committee review board to ensure compliance with ethical principles and safeguard the rights of participants. Before conducting theinterviews, all participants were provided with information about the study and were also informed about the confidentiality and anonymity of their responses.

## Data Availability

All data produced in the present work are contained in the manuscript

## References

Alcon, S., Shen, S., Wong, H.-N., Rovnaghi, C. R., Truong, L., Vedelli, J. K. H., & Anand, K. J. S. (2024). Effects of the COVID-19 Pandemic on Early Childhood Development and Mental Health: A Systematic Review and Meta-Analysis of Comparative Studies. Psychology International, 6(4), 986–1012. 10.3390/psycholint6040062

Braun, V., & Clarke, V. (2008). Using thematic analysis in psychology: Qualitative Research in Psychology: Vol 3, No 2.https://www.tandfonline.com/doi/abs/10.1191/1478088706qp063oa

Fröbel, F. (1885). The Education of Man. A. Lovell & Company.

Ginsburg, K. R., American Academy of Pediatrics Committee on Communications, & American Academy of Pediatrics Committee on Psychosocial Aspects of Child and Family Health. (2007). The importance of play in promoting healthy child development and maintaining strong parent-child bonds. Pediatrics, 119(1), 182–191. 10.1542/peds.2006-2697

Lee, K.-S., Choi, Y. Y., Kim, Y. S., Kim, Y., Kim, M.-H., & Lee, N. (2024). Association between the COVID-19 pandemic and childhood development aged 30 to 36 months in South Korea, based on the National health screening program for infants and children database. BMC Public Health, 24, 989. 10.1186/s12889-024-18361-9

Loades, M. E., Chatburn, E., Higson-Sweeney, N., Reynolds, S., Shafran, R., Brigden, A., Linney, C., McManus, M. N., Borwick, C., & Crawley, E. (2020). Rapid Systematic Review: The Impact of Social Isolation and Loneliness on the Mental Health of Children and Adolescents in the Context of COVID-19. Journal of the American Academy of Child and Adolescent Psychiatry, 59(11), 1218-1239.e3. 10.1016/j.jaac.2020.05.009

McIsaac, J.-L. D., Cummings, R., MacQuarrie, M., Lamptey, D.-L., Harley, J., Rossiter, M. D., Janus, M., & Turner, J. (2024). Shifting play experiences during the COVID-19 pandemic: Family responses to pandemic restrictions. European Early Childhood Education Research Journal. https://www.tandfonline.com/doi/abs/10.1080/1350293X.2023.2227370

Ng, C. S. M., & Ng, S. S. L. (2022). Impact of the COVID-19 pandemic on children’s mental health: A systematic review. Frontiers in Psychiatry, 13. 10.3389/fpsyt.2022.975936

Paterson, D. C., Ramage, K., Moore, S. A., Riazi, N., Tremblay, M. S., & Faulkner, G. (2021). Exploring the impact of COVID-19 on the movement behaviors of children and youth: A scoping review of evidence after the first year. Journal of Sport and Health Science, 10(6), 675–689. 10.1016/j.jshs.2021.07.001

Piaget, J. (1951). Play, Dreams and Imitation in Childhood. Psychology Press.

Singh, S., Roy, D., Sinha, K., Parveen, S., Sharma, G., & Joshi, G. (2020). Impact ofCOVID-19 and lockdown on mental health of children and adolescents: Anarrative review with recommendations. Psychiatry Research, 293, 113429. 10.1016/j.psychres.2020.113429

Spencer, H. (1870). The Principles of Psychology. Williams and Norgate.

UNICEF. (2022). COVID-19 and children. UNICEF DATA. https://data.unicef.org/covid-19-and-children/

Vygotsky, L. S., & Cole, M. (1978). Mind in Society: Development of Higher Psychological Processes. Harvard University Press.

Yogman, M., Garner, A., Hutchinson, J., Hirsh-Pasek, K., Golinkoff, R. M., Committee on Psychosocial Aspects of Child and Family Health, & Council on Communications Andmedia. (2018). The Power of Play: A Pediatric Role in Enhancing Development in Young Children. Pediatrics, 142(3), e20182058. 10.1542/peds.2018-2058

